# VR-based Gamma Sensory Stimulation: A feasibility study

**DOI:** 10.1101/2025.01.21.25320913

**Authors:** C. Reis, H. Azizollahi, G. Headley, S. Navarro, S. Hanslmayr, A. Clouter, T Zanto, R. Certain

**Affiliations:** Clarity Health Technologies, Inc., DE, USA; Centre for Neurotechnology, School of Psychology and Neuroscience, University of Glasgow, Glasgow, United Kingdom; Department of Psychology, Nottingham Trent University, Nottingham, United Kingdom; Department of Neurology, University of California-San Francisco, San Francisco, CA, USA; Neuroscape, University of California-San Francisco, San Francisco, CA, USA

## Abstract

Alzheimer’s disease (AD) presents a critical global health challenge, with current therapies offering limited efficacy and safety in halting disease progression. Gamma sensory stimulation (GSS) has emerged as a promising non-invasive neuromodulation technique that enhances gamma neural synchrony, potentially counteracting AD pathology by reducing neuroinflammation, promoting glymphatic clearance, and improving synaptic plasticity. However, existing GSS delivery methods rely on simplistic sensory stimuli that lack user engagement, creating adherence barriers and limiting the full therapeutic potential of GSS.

This pilot study assessed the feasibility, safety, and efficacy of delivering GSS via virtual reality (VR), combining EEG recordings to monitor neural activity and a digital questionnaire to evaluate safety and tolerability. Sixteen cognitively healthy older adults participated in three experiments designed to assess modulation of gamma activity in response to 40Hz stimuli. EEG recordings confirmed that unimodal auditory and visual stimuli reliably induced gamma oscillations in their respective cortical regions, while multimodal stimulation enhanced both gamma power and inter-trial phase coherence, demonstrating robust modulation of gamma activity. Notably, significant gamma activity modulation was also achieved when diversified multimedia content was modulated at 40Hz and integrated into a cognitive task, highlighting the flexibility and effectiveness of this approach.

The findings of this study validate VR as a scalable tool for delivering engaging and cognitively relevant GSS, paving the way for personalized therapies that maximize adherence and therapeutic outcomes. By integrating interactive elements, VR-based GSS may uniquely target memory-related neural networks, offering a novel approach to mitigate neurotoxicity and cognitive decline in AD.

## Introduction

Alzheimer’s disease (AD) is the leading cause of dementia worldwide, carrying a profound personal and economic burden that affects millions of families and communities (Tahami Monfared et al., 2022). Nevertheless, despite decades of research, no therapy has yet been developed that can safely provide meaningful clinical benefits while altering the progression of the disease for the vast and diverse population living with AD.

In AD, the characteristic symptoms of progressive memory loss and reduced ability to perform everyday tasks stem from multiple complex pathophysiological markers. These include the accumulation of amyloid-beta plaques and tau neurofibrillary tangles, dysregulated glial cell activity, impaired glymphatic clearance, and diminished synaptic plasticity (Crews & Masliah, 2010; Singh, 2022; Tarasoff-Conway et al., 2015). Current therapeutic strategies often focus on targeting a single pathological hallmark of AD, such as amyloid-beta accumulation or neurotransmitter imbalances, without addressing the multifactorial nature of the disease. As a result, these approaches have had limited success in modifying disease progression or restoring cognitive function in a safe and effective manner. For instance, pharmacological solutions such as cholinesterase inhibitors and NMDA receptor antagonists, which have been the standard of care for decades, provide only temporary relief from cognitive symptoms and fail to address the underlying disease pathology. Similarly, recently approved intravenous drugs like Lecanemab and Donanemab, while highly effective at reducing amyloid-beta burden, offer only modest clinical benefits and are associated with significant risks, including cerebral edema and hemorrhage (Sims et al., 2023; van Dyck et al., 2023).

In light of these challenges, non-invasive neuromodulation techniques have emerged as promising candidates for safer and potentially more effective therapeutic options for AD (Benussi et al., 2022; K. T. Jones et al., 2023; Jung et al., 2024; Koch et al., 2022; Pileckyte & Soto-Faraco, 2024). Gamma Sensory Stimulation (GSS), in particular, has gained increased attention due to its potential disease-modifying effects, high safety profile, and suitability for at-home use (Chan et al., 2022; Cimenser et al., 2021; Da et al., 2024; Hajós et al., 2024). GSS employs flickering lights and/or pulsing sounds at 40 Hz to enhance neural synchrony at gamma frequencies - a type of brain rhythms which are disrupted in AD (Kurimoto et al., 2012; Mably & Colgin, 2018)and associated with cognitive processes such as sensory integration and memory (Fries, 2015; Jensen et al., 2007). When applied over prolonged periods, GSS appears capable of modulating key disease mechanisms in a safe and impactful manner. Compellingly, Phase II clinical trial data in mild-to-moderate AD patients demonstrated encouraging results, with daily GSS for one hour over six months reducing brain atrophy by 69% and preserving cognitive and functional abilities by approximately 76% in the active treatment group compared to the placebo group. Importantly, this same study highlighted GSS’s excellent safety profile, reporting no severe adverse events and only mild to moderate adverse events that resolved quickly (Hajós et al., 2024).

These promising clinical outcomes are likely driven by the multifaceted impact GSS has on pathological processes in AD, as supported by a rapidly growing body of research. At the cellular and systemic levels, preclinical and clinical studies indicate that GSS enhances glymphatic clearance and recruits glial cells—such as microglia and astrocytes - facilitating the removal of neurotoxic proteins such as amyloid-beta, restoring homeostasis, reducing neuroinflammation, and slowing down neurodegeneration (Iaccarino et al., 2016; Q. Liu et al., 2023; Martorell et al., 2019; Murdock et al., 2024; Williams et al., 2023). At the network level, GSS is thought to recover disrupted communication pathways by promoting gamma synchronization within sensory regions and across interconnected brain areas (Blanpain et al., 2024). This synchronization supports cognitive functions and mechanisms like synaptic plasticity, which are critical for learning and memory formation (Headley & Weinberger, 2011; Wang et al., 2023).

However, despite its promise, current delivery methods for GSS, aimed at being prescribed as an at-home non-invasive neuromodulation approach, face significant limitations that may hinder its full therapeutic potential. Much like in drug discovery—where dosage, timing, and molecular targets must be carefully calibrated—neuromodulation requires optimized parameters, including intensity, frequency, and target brain regions to recover behavior (Hanslmayr et al., 2019). At present, most GSS solutions rely on passive, repetitive stimuli delivered through basic technologies like Light-Emitting Diodes (LEDs) alone or in combination with audio speakers. While effective at inducing gamma neural activity, these static designs might lack the cognitive engagement necessary to maximize therapeutic efficacy and sustain adherence, especially for long-term, at-home use.

Research suggests that the magnitude and spread of neural entrainment significantly depends on the relevance and attention given to the external stimuli (Besle et al., 2011; Fries et al., 2001; Muzzio et al., 2009). Based on intracranial EEG recordings from a single patient with refractory epilepsy, Khachatryan and colleagues, indicated that modulation of gamma activity in the hippocampus—a critical region in AD—occurred only when visual GSS was paired with cognitively engaging tasks, such as mental counting or visual attention. In contrast, passive stimulation alone was insufficient to recruit this region (Khachatryan et al., 2022). Although other studies suggest that current GSS methods can evoke gamma activity in broader memory-related networks, such as the medial temporal lobe-prefrontal cortex (MTL-PFC) network and the default mode network (DMN) (Blanpain et al., 2024; Lahijanian et al., 2024), we hypothesize that these effects may be limited in scope. Simple, passive 40 Hz flickers and tones, when delivered continuously for an hour, may not robustly or consistently engage critical deep brain regions like the hippocampus, reducing the overall impact of GSS on cognitive decline. Such a scenario would potentially help explain the mixed cognitive outcomes observed in prior studies. Notably, while the previously mentioned Phase II trial showed significant improvements on the Mini-Mental State Examination (MMSE), a general screening tool, it failed to achieve meaningful gains on its primary endpoint - Alzheimer’s Disease Assessment Scale-Cognitive Subscale (ADAS-Cog), a cognitive assessment that evaluates higher-order cognitive functions such as memory and executive processing and which is largely considered the gold standard for evaluating cognitive decline in AD (Hajós et al., 2024). This discrepancy likely reflects the inability of passive GSS to effectively engage the networks underpinning these domains.

Additionally, the monotony of current GSS protocols risks fluctuations in arousal states, potentially compromising the consistency of neural entrainment (M. Jones et al., 2019) and, consequently, therapeutic outcomes. This challenge is particularly significant in at-home settings, where maintaining sustained engagement is crucial for ensuring a positive patient experience and adherence to treatment.

To address these limitations, we propose integrating 40Hz sensory stimulation into immersive and interactive virtual reality (VR) environments as an innovative strategy to enhance the clinical efficacy of GSS and promote long-term adherence. In line with this vision, the current pilot study evaluates the feasibility, safety, and tolerability of VR-based GSS in cognitively healthy older adults. Using EEG recordings, we assessed changes in gamma rhythms during three VR-based stimulation tasks and collected safety and tolerability data through a digital questionnaire. This proof-of-concept study aims to lay the groundwork for developing user-friendly and contextually relevant VR-based GSS interventions, potentially transforming AD care by providing a widely accessible and disease-modifying treatment capable of delivering meaningful clinical benefits to the growing and diverse population affected by AD.

## Methods

### Participants

This Phase I feasibility study employed a within-patient design, where all participants were exposed to both active and sham conditions during a single study session. The study was conducted at The Sequoias Portola Valley, a senior living facility in California, and adhered to ethical guidelines, including the principles of the Declaration of Helsinki and the International Conference on Harmonization Good Clinical Practice guidelines. Ethical approval was obtained from the appropriate institutional review board and local ethics committee. All participants, or their legal representatives, provided written informed consent prior to participation, and withdrawal from the study was permitted at any time without prejudice.

A total of 16 cognitively normal older adults met the eligibility criteria and completed the study, with no participant dropouts. While the initial target enrollment was 25 participants (as outlined in the clinicaltrials.com registration, NCT06234930), the study employed an adaptive design, enabling the assessment of the primary outcomes (modulation of gamma activity, safety and tolerability) at n=15 participants. Inclusion criteria required participants to have a Montreal Cognitive Assessment (MoCA) score equal or above 26, at least 8 years of formal education, fluency in English, and sufficient vision and hearing ability to engage with the study tasks. Participants were excluded if they had: received Memantine treatment in the past 30 days; initiated treatment with acetylcholinesterase inhibitors within the past 30 days; a history of seizure or epilepsy (including a family history of such conditions); a history of stroke; an active diagnosis of migraine headache; diagnosis of mild Alzheimer’s disease or related dementias, or a Geriatric Depression Scale (GDS) score greater than 8. Sufficient vision and hearing to participate in our study were determined by a calibration session where participants had to repeat sentences played to them through the VR headset and describe images displayed via the VR screen.

**Table 1.**
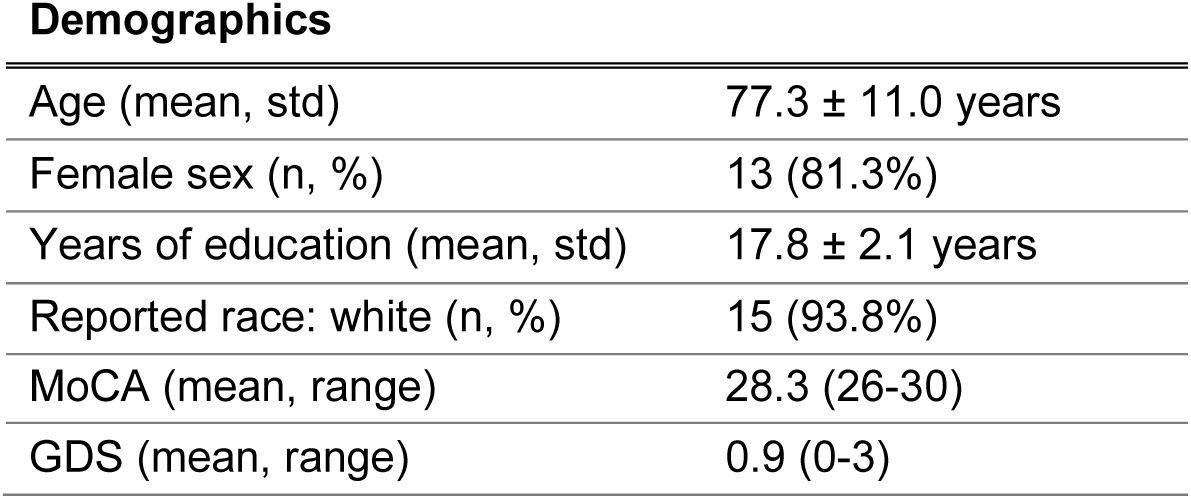
Demographics and baseline clinical characteristics of cognitively healthy older participants (n=16).

### Materials

Participants received multisensory stimulation through one of two VR headsets (Oculus Quest 2), tethered to a computer. Visual stimuli were delivered via the VR headset display, while auditory stimuli were presented through headphones (SONY MDR-EX15LP) connected to the headset.

To evaluate the device’s performance in delivering synchronized and stable audiovisual stimuli, stability tests were conducted pre- and post-experimental sessions, as it was not physically feasible to record this data during the experimental session. An Arduino Nano microcontroller was used to facilitate the audio and visual recordings of the stimulation. To capture auditory outputs from the VR headset, we interfaced the Arduino board with the headset via a 3.5mm audio jack. For visual signal measurements, a TEMT6000 photosensor was employed to detect brightness levels from the VR headset’s display.

Visual and auditory stimuli were varied depending on the experiment, but the subtended flickered visual field was kept constant across the whole study, comprehending a 54.5° diagonal visual field (48.7° horizontal, 28.6° vertical). Videos and sounds were loaded to the device with their brightness and volume premodulated by a 40Hz sine wave. The overall VR headset sound volume level of was adjusted to the comfort level of each participant. All auditory stimuli were normalized to 0.0 dB maximum and had a 100 ms linear amplitude fade-in/ fade-out to avoid a startle effect.

Throughout the experiment, EEG recordings were collected using the 64 channel BioSemi ActiveTwo system (BioSemi, Amsterdam, the Netherlands). Electrode positions followed the 64 standard equidistant BioSemi sites; however, the five most frontal electrodes (AF7, FP1, FPz, FP2, AF8) were removed to ensure participant comfort, as the top edge of the VR headset overlapped with these positions. EOG generated from eye movements and blinks were recorded from two additional electrodes placed approximately 1 cm to the left of the left eye and 1 cm to the right of the right eye. Data was digitized using the BioSemi ActiView software, with a sampling rate of 2048 Hz. Offline analyses were performed with MNE-Python (Gramfort et al., 2013).

### Experimental Tasks

#### Testing Changes in Gamma Activity in Response to 40Hz stimuli

To evaluate the feasibility of using a VR-based gamma sensory stimulation protocol to drive neural activity at gamma frequencies, we designed three complementary experiments. Experiment 1 focused on the isolated effects of unimodal auditory and visual stimuli modulated at 40Hz. Experiment 2 aimed to disentangle gamma stimulus-driven gamma synchronization from endogenous sensory processing by comparing neural responses to 40Hz-modulated multimedia content against an unmodulated sham condition. Finally, Experiment 3 investigated whether modulation of gamma activity could be achieved with more complex, multimodal audiovisual stimuli delivered during a cognitively engaging associative memory task. Together, these experiments systematically assessed the efficacy and flexibility of VR-based gamma sensory stimulation.

#### Experiment 1 - Passive Unimodal 40Hz VR-based Stimulation

This experiment aimed to assess the feasibility of entraining neural activity to gamma rhythms using distinct visual and auditory stimuli presented in a VR environment. A total of 100 trials were conducted, evenly divided between visual and auditory conditions, with each trial lasting 3 seconds and separated by a jittered inter-trial interval of 3.5 to 4 seconds. During the inter-trial interval, a black screen with a white fixation cross at the center was displayed to maintain participant focus. Visual stimulation, with brightness sinusoidally modulated between 0% and 100% at 40Hz (pixel brightness range: 0–255), consisted of 50 trials split into two categories: 25 trials of a plain white rectangle with an average pixel brightness of 255 (referred to as the black and white video), and 25 trials of neutral-valence videos, repeated five times each, with an average brightness of 94.41 (referred to as random bright videos). Auditory stimulation also consisted of 50 trials, comprising 25 trials of isochronous 40Hz click trains characterized by waveforms with 0 ms rise/fall times and 1 ms plateau durations (similar to prior work by (Arnal et al., 2019), and referred to as clicks); and 25 trials of amplitude-modulated pure tones at 40Hz, featuring five distinct carrier frequencies evenly spaced between 196 Hz and 1000 Hz, each repeated five times (referred to as pure tones). This experimental design systematically tested the ability of controlled unimodal conditions to drive gamma neural activity, allowing for a comparison of simple and complex sensory stimuli in both visual and auditory modalities.

#### Experiment 2 - Passive Multimodal 40Hz VR-based Stimulation

This short experiment was aimed at validating that the increased gamma neural activity observed during 40Hz modulated multimedia (audiovisual) content, was driven by the repetitive external stimuli rather than by endogenous gamma activity associated with sensory processing of the multimedia content itself. To this end, participants passively observed and listened to the same video under two conditions: with its brightness and sound volume simultaneously modulated at 40Hz (active condition) and without any modulation (sham condition). The visual content of the movie had an average pixel brightness of 177.5, while the auditory stimuli consisted of an amplitude-modulated 196Hz pure tone. Each condition included 15 trials, with each trial lasting 5 seconds separated by a 7-second inter-trial interval. During the inter-trial interval, an immersive black screen was displayed with a white fixation cross at the center. Active and sham trials were randomly displayed to avoid an order effect.

#### Experiment 3 - Active Multimodal 40Hz VR-based Stimulation

In Experiment 3, we aimed to determine whether loosely controlled multimedia content—defined here as audiovisual stimuli with naturally varying characteristics, such as brightness, auditory frequency range, and complexity—modulated at 40Hz and integrated into an associative memory task could induce a significant increase in gamma neural activity compared to the sham condition.

To achieve this, we adapted a video-sound associative memory task previously used in sensory stimulation research (Clouter et al., 2017; Wang et al., 2018, 2023). Participants were instructed to focus on and memorize video-sound pairs that were either modulated at 40Hz and lasted 3 seconds (stimulation condition) or unmodulated and lasted 1.5 seconds (sham condition) - a strategy used to balance the amount of information conveyed to the participant across the two conditions. The videos used were of neutral valence and varied significantly in their average brightness (60.78 - 182.93). To ensure comparability, the distribution of the average pixel brightness of videos used in the 40Hz stimulation condition was matched to the average pixel brightness of the videos used in the sham condition (respective means and standard deviation: 105.05 ± 23.45 and 107.4 ± 23.57, t-test=-0.598, p=0.551). Auditory stimuli were drawn from four distinct categories—piano keys, orchestra, acoustic guitar, and electric guitar— with an equal number of sound types used in both the active and sham conditions. The sounds’ spectral entropy, a normalized metric where 0 indicates minimal complexity and 1 indicates high complexity, ranged from 0.19 to 0.67, reflecting the diverse sound characteristics used in the task.

Immediately following each video-sound pair association, participants verbally rated how well the video matched the sound on a 5-point scale, where 1 indicated “very poorly” and 5 indicated “very well”. Following their response, an inter-trial interval of 1–3 seconds occurred, during which a white fixation cross was displayed. After every 12 trials in the encoding phase, participants completed a distractor task which involved a visual search task (counting specific target objects among distractors on a table) that lasted 1 minute before entering the recall phase. During the recall phase one of the previously played sounds was played again and three images were displayed simultaneously. The participant was asked to selected from 1 to 3 the image that corresponded to the video previously played together with the sound. Of the three options, one was the correct match, one was a foil (an image never shown during the encoding phase), and one was a distractor (an image presented earlier but not associated with the current sound).

By assessing neural responses under these more naturalistic and engaging multimodal conditions, this experiment was design to evaluate the robustness of VR-based stimulation for enhancing gamma oscillatory activity during a cognitively loaded task.

#### Testing Safety and Tolerability

After completing all experimental tasks and removing the VR headset and EEG cap, participants completed a digital questionnaire to evaluate the safety and tolerability of the VR-based GSS protocol. The questionnaire assessed potential side effects, including headaches, eye pain, disorientation, and nausea, as well as measures of tolerability, such as comfort, enjoyment, and the perceived intensity of the VR experience. Participants rated the severity of side effects on a scale from 0 to 3, where 0 indicated “not at all,” 1 indicated “mild,” 2 indicated “moderate,” and 3 indicated “severe”. To measure participants’ comfort, enjoyment, and tolerance of various aspects of the protocol and the VR headset, responses were recorded using a 7-point Likert scale. Here, 1 represented the lowest level (e.g., “not at all”), and 7 represented the highest level (e.g., “extremely”).

#### Testing Stability of VR-Based Stimuli Delivery System

To evaluate the performance of the off-the-shelf VR devices used in our experiments (n=2, alternated between consecutive participants), we conducted stimulation stability tests and quantified the stability of our 40Hz visual and auditory stimulation, as well as the synchronization between audiovisual stimuli (as required in Experiments 2 and 3). These tests were performed twice for each participant: once before the recording session began and again after the session ended.

For these tests, an Arduino device was used to collect two types of signals: the voltage fluctuations corresponding to the sound output from the VR headset and the brightness of the VR display using a photodiode. The experiment involved displaying the same movie used in Experiment 2, with its brightness and sound amplitude modulated at 40 Hz with 100% depth. Data were recorded across 25 trials, each lasting 1 second with 3 seconds between trials.

Results across all participants and trials indicated stable delivery of stimuli. For visual stimuli, the average peak frequency in the power spectral density was 39.95 ± 0.67 Hz, while for auditory stimuli, it was 40.43 ± 0.52 Hz. The delay between audio and visual stimulus onset was -8.8 ± 5.0 ms, with negative values indicating that the visual stimulus preceded the audio stimulus. Phase consistency, measured as phase locking value (PLV) between a simulated 40Hz sinusoidal wave an individual trials, was 0.65 ± 0.16 for visual stimuli and 0.92 ± 0.03 for auditory stimuli. PLV between audio and visual data was on average 0.68 ± 0.16. Importantly, none of these metrics varied significantly between pre- and post-session tests (p=0.503 and p= 0.486 for audio and visual PLV, respectively and p= 0.137 for PLV between auditory and visual stimuli data).

Although real-time device performance data could not be collected during experimental sessions due to form factor limitations, these stability tests confirm that the VR-based stimuli maintained an average periodicity at the frequency of interest, i.e., 40Hz, which we expect to have been maintained across the study as these metrics did not vary significantly between pre and post session. Together these findings validate the reliability of our VR-based software system in generating periodic 40Hz audiovisual stimuli, ensuring the integrity of the experimental protocol.

#### Metrics to evaluate neural responses to sensory stimuli

##### EEG Preprocessing

All participants’ data underwent identical preprocessing steps. The raw signals were initially subjected to a notch filter at 60 Hz and 120 Hz using a finite impulse response (FIR) filter to eliminate power line noise and its harmonics. Subsequently, a band-pass FIR filter was applied with cutoff frequencies set at 1 Hz and 100 Hz to retain the relevant EEG frequency range while removing low-frequency drifts and high-frequency noise. The data were then resampled to a sampling frequency of 1024 Hz to enhance the computational efficiency. To standardize the reference across all recordings, the EEG signals were re-referenced to average reference. Independent Component Analysis (ICA) was then applied to the data, and components were rejected if they represented muscle artifacts, eye movements, heart activity, channel noise, and other types of artifacts. To quantify stimulus-evoked responses to auditory and visual stimuli at the sensor level, we selected among parieto-occipital electrodes and frontocentral electrodes for visual and auditory stimulation respectively, the four channels that underwent the largest modulation at gamma frequencies across all stimulation trials and participants. As a result, electrodes FCz, C2, C1, and Cz were used to assess neural responses to auditory stimuli, and electrodes Oz, PO3, PO4, and POz were used to address neural response to visual stimulation. For combined visual/auditory stimuli, as well as the sham condition, both sets of electrodes were used (n=8, FCz, C2, C1, Cz, Oz, PO3, PO4, and POz).

##### Power spectral density (PSD)

Power spectral density estimates were obtained using Welch’s method as implemented in the MNE-Python package. For each epoch, the selected channels and specified time interval were segmented into overlapping windows and multiplied by a tapering function prior to computing the Fourier transform. The resulting periodograms were averaged to yield stable PSD estimates over the 1–100 Hz frequency range. This approach reduces spectral variance, facilitating a more robust characterization of the underlying neural dynamics.

##### Wavelet Power analysis

Gamma power was computed by convolving each EEG signal with a family of Morlet wavelets across (38 - 42 Hz). Specifically, the Morlet transform decomposes the signal into time-frequency space, and the squared magnitude of the resulting complex coefficients corresponds to the instantaneous power at each time point and frequency. These instantaneous power values are then averaged or integrated within the relevant time windows (e.g., baseline vs. stimulation intervals) to yield a final power estimate for each condition and gamma frequency band of interest. To evaluate stimulus-related changes in power at 40 Hz, we compared power values between a baseline period and the stimulus presentation period. To account for the varying active and inter-trial intervals across the experimental tasks, different baseline windows were defined: -1 to 0 seconds relative to stimulus onset for Experiment 1, -4.5 to -0.75 seconds for Experiment 2, and -2 to 0 seconds for Experiment 3. Similarlly, the stimulus presentation period was defined as 0.75 to 2.5 seconds for Experiment 1, 0.75 to 4.5 seconds for Experiment 2, and 0.75 to 2.5 seconds for Experiment 3. We initiated the stimulus presentation analysis window at 0.75 seconds post-stimulus onset. This delay was implemented to minimize contamination from early event-related potentials (ERPs), thereby isolating sustained neural responses from transient, stimulus-locked activity. The resulting power values were averaged across the selected channels for each condition.

##### Inter-Trial Phase Coherence (ITPC)

ITPC was computed using the MNE-Python package which implemented Morlet wavelet transformations. The analytical procedure involved the convolution of the selected channels (mentioned above) and specified time interval epoch with complex Morlet wavelets to derive the time-frequency representation, yielding both amplitude and phase information at each temporal and spectral point. An ITPC value was then computed by subsequently averaging the phase components across trials. The ITPC at a given frequency *f* and time point *t* was defined as:

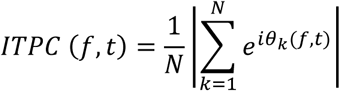

Where *f* is frequency of interest, *t* is time point of interest, *N* is number of trials, θ_*k*_(*f*, *t*) is phase of the signal in the *k* − *t*ℎ trial at frequency *f* and time *t* and |. | denotes magnitude of the resultant vector.

This methodological approach provides a good measure of the extent to which the oscillatory neural activity is phase locked to the experimental stimuli and thus provides an insight into the time course of the neural synchronization. To evaluate stimulus-related changes in phase coherence, we compared ITPC values between a baseline period (−1 to 0 seconds relative to stimulus onset for experiment 1, -4.5 to -0.75 seconds for experiment 2 and -2 to 0 seconds for experiment 3) and the stimulus presentation period (0.75 to 2.5 seconds and 0.75 to 4.5 for experiment 1 and 3 an experiment 2, respectively). The resulting values were averaged across the frequency range of interest (38-42 Hz) and across the selected channels for each condition.

##### Fold Change

Similar to prior work (Blanpain et al., 2024; Duecker et al., 2021), to facilitate the direct interpretation of our results, significant effects of VR-based GSS on neural activity were quantified using the fold change (FC) in power and inter-trial phase coherence (ITPC) at the stimulation frequency. The Fold Change (FC) was calculated as the ratio of ITPC or power values during the stimulation period to the corresponding values in the baseline period, as shown in the equation below. Subsequently, we subtracted 1 from this ratio yielding a unitless measure centered around zero, where positive values indicate an increase from baseline, and negative values indicate a decrease from baseline.

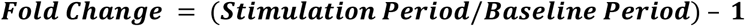

##### Time Frequency Analysis

Time-frequency representations of power and inter-trial phase coherence (ITPC) were computed using the Stockwell transform (S-transform) as implemented in the MNE-Python package. The S-transform was applied across a frequency range of 1-85 Hz, with a width parameter of 0.5 to balance temporal and frequency resolution. For both power and ITPC analyses, the temporal resolution was reduced by a factor of 3 (decim=3) to enhance computational efficiency. Power spectrograms were calculated as the squared magnitude of the S-transform coefficients and then converted to decibels (dB) using a log-ratio transformation relative to a baseline period of -4.5 to -0.75 seconds in Experiment 2. ITPC was computed by first normalizing the complex S-transform coefficients to unit magnitude for each trial and then averaging these normalized coefficients across trials. This resulted in a measure of phase consistency ranging from 0 to 1.

##### Source level analysis

In this study, we applied the Dynamic Imaging of Coherent Sources (DICS) beamformer method (as implemented in MNE-Python) to localize induced gamma-band activity (38–42 Hz) of unimodal stimuli (Experiment 1) to cortical regions. First, we loaded epoched EEG data from −1 to 3 s relative to stimulus onset and co-registered these data to the fsaverage template brain (Freesurfer) by computing a forward solution, ensuring consistency across participants. We then computed cross-spectral density (CSD) matrices using Morlet wavelets (decimated by a factor of 20), and optimized the DICS filter parameters by conducting an exhaustive search over multiple regularization values combined with five-fold cross-validation. Using the optimal filter, we estimated source power during both the baseline (−1 to 0 s) and activation (0.75 to 2.5 s) periods for each epoch. We then calculated the log-ratio (in decibels) of the activation power relative to baseline power and morphed these estimates to the fsaverage brain. Finally, individual source estimates were averaged to generate a grand-average source map, which served as the basis for visualizing the spatial distribution of gamma-band activity.

##### Statistical Analysis

In this study, we employed a range of statistical tests to test the significance of change in gamma power and phase alignment from baseline, within and across conditions. We first assessed the normality of each dataset using the Shapiro-Wilk test. For paired comparisons, such as baseline versus stimulation, we used a paired t-test (or one-sample t-test if appropriate) if the data were normally distributed and a Wilcoxon signed-rank test if they were not, to calculate p-values. For comparisons between independent groups, such as two different experimental conditions, we again assessed normality and then used either an independent-samples t-test for normally distributed data or a Mann-Whitney U test for non-normally distributed data. When evaluating a single condition against a theoretical value, like a fold change of 1, we used a one-sample t-test or a Wilcoxon signed-rank test, depending on the data’s normality. Significant differences are indicated in the figures with their corresponding p-values, using asterisks to denote the level of significance (* for p < 0.05, ** for p < 0.01, *** for p < 0.001).

## Results

### VR-based Gamma Sensory Stimulation is safe and well-tolerated

Safety and tolerability assessments from the digital questionnaires completed by all 16 participants at the end of the experimental session revealed that our VR-based gamma sensory stimulation protocol was well-received, with no severe adverse events reported (see Table 2). Notably, no participants experienced motion sickness or tinnitus, underscoring the safety of integrating sensory stimulation protocols with VR technology.

**Table 2.** Results from the Safety Questionnaire completed by 16 cogntively healthy older participants.

| **Adverse Events**<br>(n=16) | **Null**<br>n. participants<br>(% participants) | **Mild**<br>n. participants<br>(% participants) | **Moderate**<br>n. participants<br>(% participants) |
| --- | --- | --- | --- |
| General discomfort | 7 (43.7%) | 8 (50%) | 1 (6.3%) |
| Headache | 12 (75%) | 4 (25%) | - |
| Tinnitus | 16 (100%) | - | - |
| Eye pain | 14 (87.4%) | 1 (6.3%) | 1 (6.3%) |
| Disorientation | 15 (93.7%) | 1 (6.3%) | - |
| Fatigue | 5 (31.2%) | 7 (43.8%) | 4 (25%) |
| Nausea | 16 (100%) | - | - |
| Blurred vision | 10 (62.5%) | 6 (37.5%) | - |

Tolerability results further highlighted the acceptability of the protocol. Most participants found the headset comfortable, with 68.8% rating it 5 or higher on a 7-point scale (1 = “not at all comfortable” and 7 = “extremely comfortable”), and the VR experience was rated as enjoyable, with no participant giving a score below 4 (4 = “neutral enjoyment”). Additionally, 93.8% of participants found the VR environment manageable and non-overwhelming, rating it 2 or lower on a 7-point scale (1 = “not at all overwhelming” and 7 = “extremely overwhelming”), with only one participant score of 4.

These findings demonstrate that our VR-based GSS approach provided a safe and balanced immersive experience suggesting its viability for extended use. Although this study primarily assessed feasibility with added equipment (EEG cap and VR headset) and multiple experimental tasks instead of a complete therapeutic protocol, these preliminary results provide a solid foundation for the development of a scalable, safe, and effective therapeutic application.

### VR-based Gamma Sensory Stimulation leads to significant modulation of gamma activity

Experiment 1 assessed the ability of VR technology to drive neural activity at gamma rhythms by separately presenting distinct visual and auditory stimuli. Both sensor-level (Figure 1A, B, and C) and source-level (Figure 1D) analyses confirmed that our VR-based GSS protocol was capable of effectively enhancing gamma oscillatory activity. This enhancement was evidenced by the clear spectral peak at 40Hz (dashed vertical line) in the average power spectral density curves during active stimulation, and its absence during baseline periods (Figure 1A and 1C).

**Figure 1.**
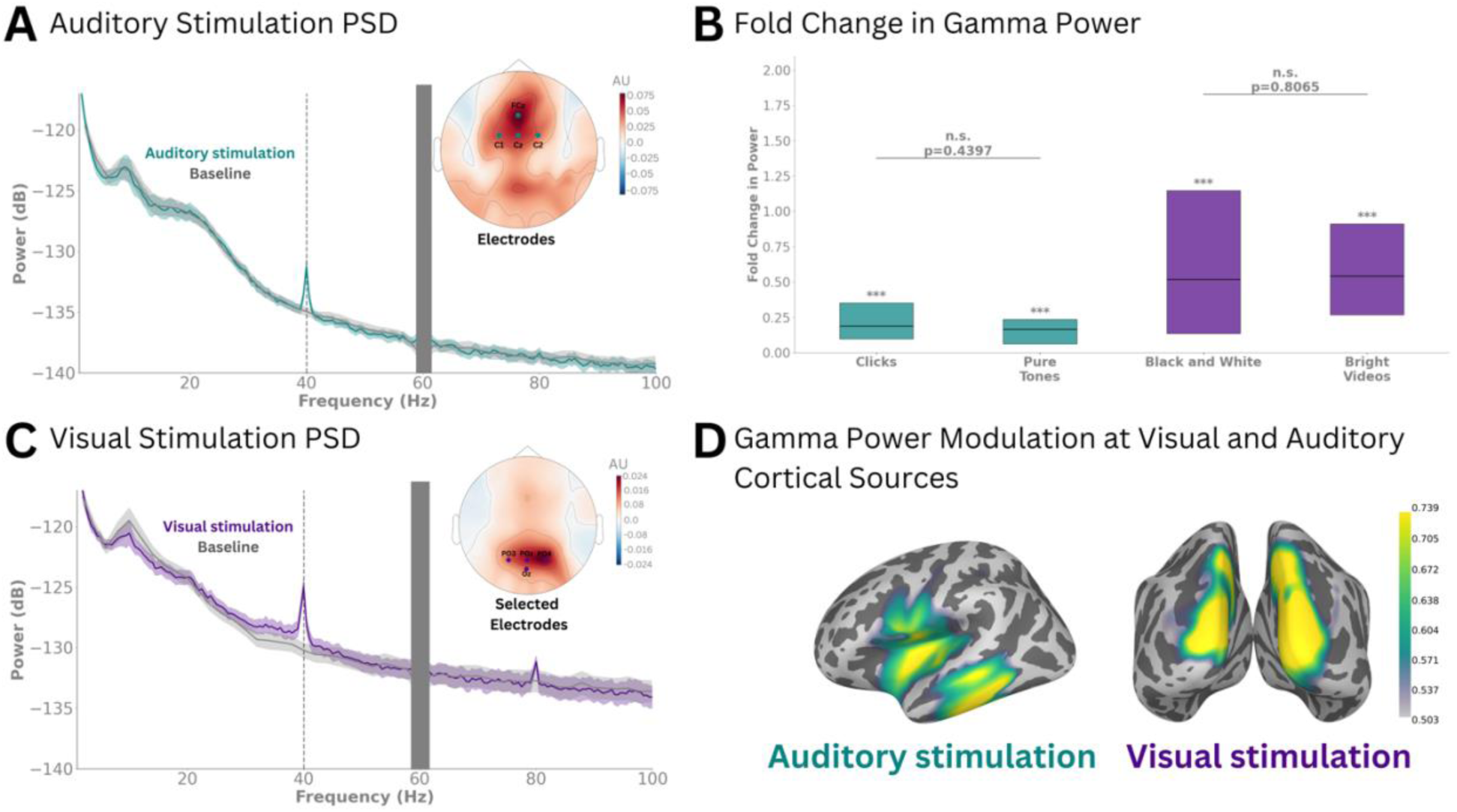
Gamma power modulation during Auditory and Visual VR-based 40Hz stimulation. **(A)** Power spectral density analysis comparing auditory stimulus (green line) and baseline periods across frontocentral channels (Cz, C2, C1, and FCz). The plot shows grand average power spectra during auditory stimulation (green line) and baseline periods (gray line). Shaded areas represent the standard error of the mean around each period. The topo map depicts the spatial distribution of power in gamma band (38-42 Hz) at the sensor level using baseline corrected log power ratio. The colour scale represents the relative change in power from baseline, where darker blue indicates lower power and red indicates higher power. (**B)** Comparison of gamma band (38-42 Hz) power changes across different sensory stimulation conditions. The plot shows fold changes in power between stimulation and baseline periods and between stimulation conditions. Box plots represent power changes for auditory conditions (green: Clicks, Pure Tones) and visual conditions (purple: Black and White, Bright Videos). The data is presented as box plots where the horizontal line represents the median, boxes show the interquartile range, and individual significance is indicated by asterisks (*p < 0.05, **p < 0.01, ***p < 0.001). **(C)** Same as (A) but for visual stimulation and across parietooccipital channels (POz, PO3, PO4, Oz). **(D)** Cortical source localization of gamma power during auditory (green) and visual (purple) stimulation, highlighting modality-specific cortical gamma modulation. The figure shows DICS beamformer source reconstruction of gamma band (40 Hz) activity, comparing stimulation to baseline periods. Color intensity represents the log-transformed power ratio (dB) between stimulation and baseline periods.

Further analysis revealed that all stimuli modulated at 40Hz - whether simple or complex, auditory or visual - elicited a significant increase in gamma power from baseline. At the group level, fold-changes (FC) in gamma power were observed at sensor locations relevant to the modality: Oz, PO3, PO4, and POz for visual stimuli, and Cz, C2, C1, and FCz for auditory stimuli. Specifically, auditory stimuli showed significant gamma power increases for both 40Hz click trains and pure tones amplitude-modulated at 40Hz, with no significant difference in the magnitude of their effects (FCclicks = 0.19, p < 0.001; FCpure tones = 0.17; p < 0.001; difference between FCclicks and FCpure tones = 0.02, p = 0.4397). Similarly, visual stimuli demonstrated significant gamma power increases for both black-and-white videos and random bright videos (FCblack-and-white videos = 0.52, p<0.001; FC random bright videos = 0.54, p<0.001), with no significant difference in gamma power increases between the two types of visual stimuli (difference between FCblack-and-white and FCrandom white videos = -0.02, p = 0.8065).

Source-level analysis further localized the increase in gamma activity to the primary auditory and visual cortical areas for the auditory and visual stimuli, respectively (Figure 2D). This finding confirms the specificity of the neural response observed in previous analyses and rules out contributions from muscular activity (e.g., eye squinting due to the flicker or neck tension from the headset weight), which would typically be registered over the pre-central gyrus, or electrical contributions from the device or headphones, which would be predominantly registered over frontal and temporal regions respectively.

**Figure 2.**
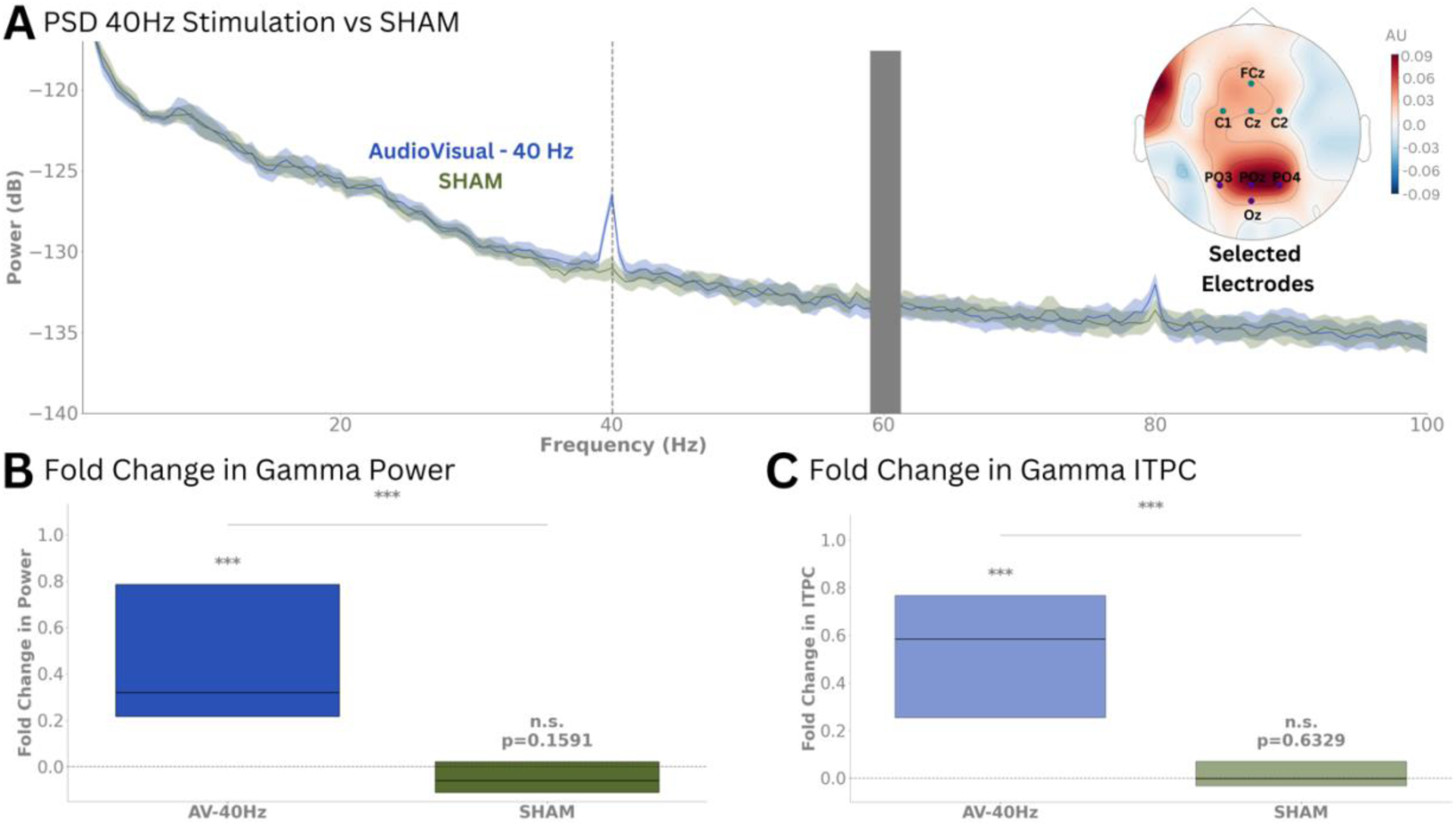
Modulation of gamma neural activity in response to audiovisual VR content modulated at 40Hz. **(A)** Group-level PSD comparison between 40 Hz audiovisual stimulation and sham conditions. The plot shows grand average power spectra (± SEM, shaded areas) across frontocentral and parietooccipital channels (FCz, C1, C2, Cz, Oz, PO3, PO4, POz). Power spectra are shown for the 40Hz stimulation condition (labelled as AudioVisual-40 Hz stimulation and shown in blue) and sham condition (labelled as SHAM and shown green). The topo map depicts the spatial distribution of power in gamma band (38-42 Hz) at the sensor level using baseline corrected log power ratio. The colour scale represents the relative change in power from baseline, where darker blue indicates lower power and red indicates higher power. (B) Fold change in gamma (38-42 Hz) power from stimulation to baseline periods during 40 Hz audiovisual stimulation (AV-40Hz, in blue) and sham conditions (SHAM, in green). **(C)** Same as (B) but for ITPC. The data is shown as box plots, where the horizontal line marks the median and the boxes outline the interquartile range. Significant differences are indicated by asterisks (*p < 0.05, **p < 0.01, ***p < 0.001).

Together, these findings underscore the ability of VR-based stimulation to reliably modulate gamma activity across diverse stimulus types, whether simple or complex, in both auditory and visual modalities.

In Experiment 2, we evaluated the feasibility of delivering audiovisual stimulation through VR technology and sought to validate that the observed gamma activity was driven by external 40Hz modulation rather than endogenous sensory processing (Bartoli et al., 2019; Duecker et al., 2021; Lachaux et al., 2005; Tallon-Baudry et al., 1998). This was achieved by comparing neural responses under two conditions: brightness and sound volume modulated at 40Hz (active condition) versus no modulation (sham condition), and by measuring neural responses as fold-change from baselinenot only in gamma power but also in ITPC at gamma frequencies.

Similar to the results obtained under unimodal conditions, multimodal 40Hz stimulation significantly enhanced gamma activity, as shown by a prominent spectral peak at 40Hz in the PSD present only for the active condition (Figure 2A). Moreover, both gamma power and ITPC increased significantly from baseline in the active condition (FC power AV-40 = 0.32, p < 0.001; FC ITPC AV-40 = 0.58, p < 0.001), while no significant changes were observed in the sham condition (FC power Sham = -0.06, p = 0.159; FC ITPC Sham = 0.00, p = 0.633) (Figures 2B and 2C).

Figure 3, depicts a single-participant analysis highlighting the spatial localization of the 40Hz stimulation at the sensor level and its temporal stability. Similarly to figure 2A, figure 3A shows a 40Hz spectral peak during stimulation but not during baseline. Additionally, the polar plots (Figure 3B) depicting the distribution of phase differences in gamma rhythmic activity (38–42Hz) across pre- and post-stimulation periods, reveal consistent phase alignment between gamma rhythmic activity across trials in frontocentral and parietoocipital electrodes. Time-frequency (Figure 3C) and ITPC plots (Figure 3D) confirm sustained gamma power increases and phase-locking during the stimulation window, supporting our group-level findings.

**Figure 3.**
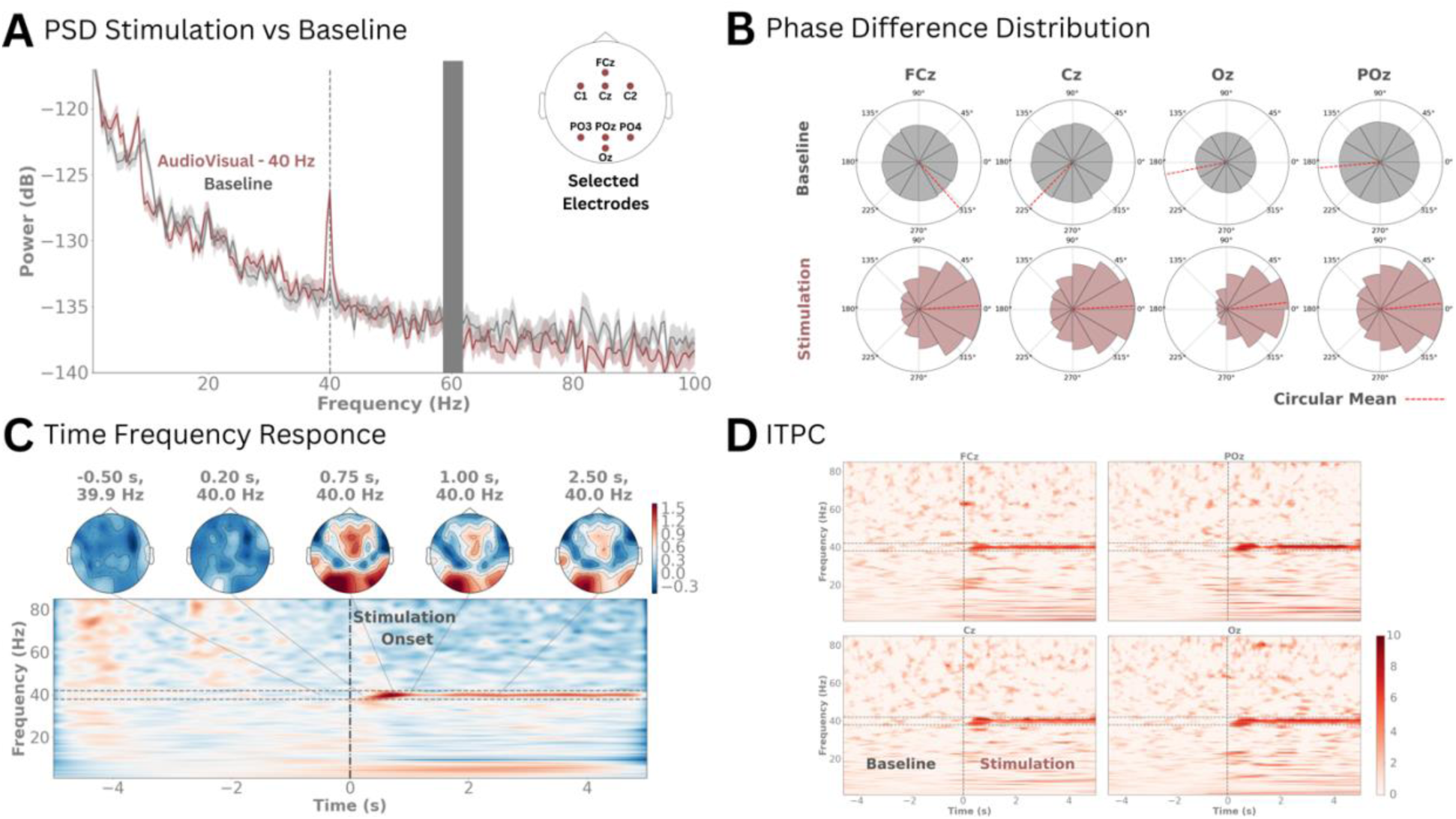
Individual subject results for 40 Hz audiovisual stimulation. **(A)** Power spectral density (PSD) comparison between 40 Hz audiovisual stimulation (red) and baseline. Shaded areas represent standard error of the mean (SEM). **(B)** Phase difference polar plots for selected electrodes (FCz, Cz, Oz, POz) comparing baseline and stimulation conditions. The polar histograms show the distribution of instantaneous phase differences across multiple trials in 12 bins from -π to π. Top row displays baseline period phase distributions (grey), while bottom row shows stimulation period phase distributions (red). Red dashed lines indicate circular means of phase angles. **(C)** Joint visualization of time-frequency power changes and corresponding topographical distributions during 40 Hz stimulation. Top row shows topographical maps of power distribution at five time points relative to stimulation onset. The topographic map uses a color scale ranging from -0.3 (blue) to 1.5 (red), representing log-ratio power changes relative to baseline, with white indicating no change (0). The main panel shows the time-frequency representation of power changes across the epoch (−5 to 5s), with horizontal dashed lines indicating the gamma band of interest (38-42 Hz) and vertical dash-dotted line marking stimulation onset (0s). **(D)** Inter-trial phase coherence during audio-visual stimulation for four selected EEG channels (FCz, Cz, Oz, POz). Time-frequency representations show inter-trial phase coherence (ITPC) computed using Stockwell transform (1-85 Hz). The data is baseline corrected and displayed in z-scores using a red colormap (0 to 10). Gray dashed lines indicate the frequency band of interest (38-42 Hz). Higher ITPC values (darker red) represent stronger phase consistency across trials at specific time-frequency points.

Altogether, these findings demonstrate that VR-based 40Hz multisensory stimulation effectively induces robust oscillatory activity at gamma frequencies, confirming its efficacy in multimodal conditions.

In Experiment 3, we assessed whether loosely controlled multimedia content, modulated at 40Hz and integrated into a video-sound associative memory task, could effectively drive gamma neural activity. As shown in Figure 4, both gamma power and ITPC significantly increased during the active 40Hz modulation condition compared to baseline (FC power AV-40 = 0.17, *p* < 0.001; FC ITPC AV-40 = 0.29, *p* < 0.001), with no significant changes observed in the sham condition for either metric (FC power Sham = -0.03, *p* = 0.579; FC ITPC Sham = 0.0, *p* = 0.873). Moreover, the increases in gamma power and ITPC during the active condition were significantly greater than those in the sham condition (*p* < 0.001). A secondary aim of this task consisted of testing whether content flickered at 40Hz versus non-flickered content (sham condition) during the encoding phase of the associative memory task led to higher levels of recall accuracy. We found no significant differences in the percentage of correct recall trials between the two conditions (mean ± std accuracy for the active and sham conditions: 64% ± 10% and 66% ± 11%, respectively; *p =* 0.372). Importantly, the these do not rule out the possibility that applying these protocols longitudinally could produce cognitive and neuroprotective benefits for AD patients, as explored further in the discussion section.

**Figure 4.**
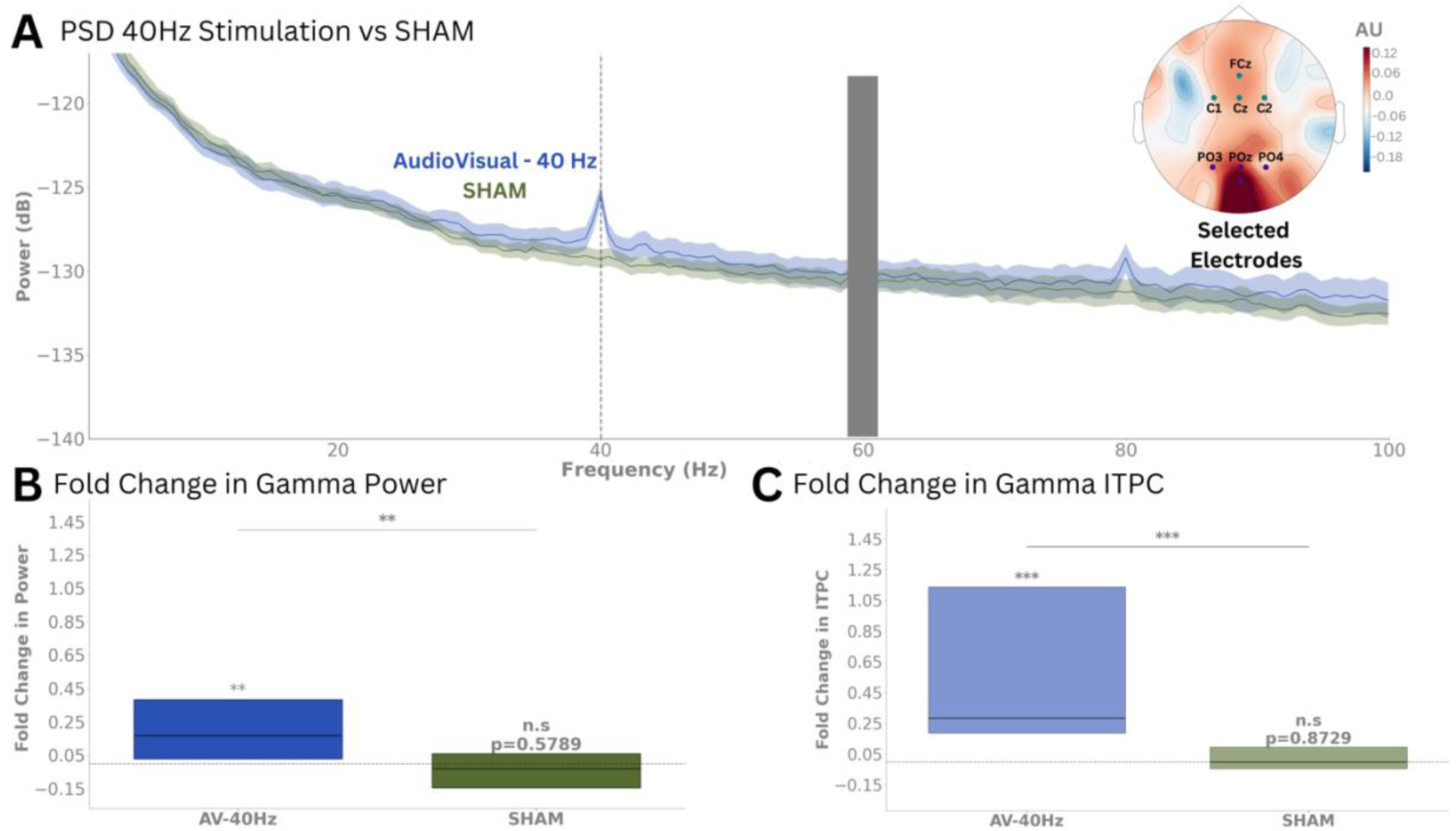
Modulation of gamma neural activity in response to task-based VR Gamma Sensory Stimulation. **(A)** Group-level PSD comparison between 40 Hz audiovisual stimulation and sham conditions. The plot displays grand average power spectra (± SEM, shaded regions) across selected channels (FCz, C1, C2, Cz, Oz, PO3, PO4, POz). Power spectra are illustrated for the 40 Hz stimulation condition (labeled as AudioVisual-40 Hz stimulation and represented in blue) and the sham condition (labeled as SHAM and depicted in green). The topo map depicts the spatial distribution of power in gamma band (38-42 Hz) at the sensor level using baseline corrected log power ratio. The colour scale represents the relative change in power from baseline, where darker blue indicates lower power and red indicates higher power. **(B)** Fold change in gamma (38 - 42 Hz) power. **(C)** Fold change in gamma (38-42 Hz) ITPC (38 - 42 Hz) for selected (FCz, C1, C2, Cz, Oz, PO3, PO4, POz) channels. The plot shows fold changes in power and ITPC between stimulation and baseline periods for 40 Hz audiovisual stimulation (blue) and sham conditions (green). The data is depicted by box plots where the horizontal line represents the median, boxes show the interquartile range, and individual significance is indicated by asterisks (*p < 0.05, **p < 0.01, ***p < 0.001).

These findings demonstrate that a reliable increase in gamma activity can be achieved even with multimedia content featuring varying brightness, auditory complexity, and naturalistic characteristics. This marks an important step toward developing flexible VR-based therapies that maintain effective modulation of gamma brain rhythms while fostering greater patient engagement and adherence through more immersive and cognitively-relevant GSS protocols.

## Discussion

This feasibility study demonstrates that virtual reality (VR) technology can effectively deliver gamma sensory stimulation (GSS). In a cohort of 16 cognitively healthy older adults, we showed that 40Hz-modulated audiovisual content, delivered under both passive and cognitively demanding conditions, successfully and safely evoked gamma oscillatory activity. These findings establish a strong proof of concept for leveraging VR technology into engaging and cognitively relevant GSS interventions aimed at enhancing both the therapeutic efficacy and adherence of current GSS approaches.

From a safety perspective, the absence of severe adverse events aligns with prior findings from a six-month trial investigating self-administered daily GSS in mild to moderate AD (Hajós et al., 2024). In the current study, the most frequently reported side effects, including mild headaches, general discomfort, and fatigue, were transient and are most likely attributable to the experimental setup, which involved multiple tasks and the use of an EEG cap, headphones, and a VR headset. This suggests that these effects are unlikely to stem from the neuromodulation approach itself and emphasizes the need for longitudinal trials with less burdensome setups to further assess tolerance and patient adherence to VR-based GSS. Tinnitus, a phantom auditory perception characterized by a persistent ringing or buzzing in the ears, has been reported as a side effect in previous studies of GSS. It occurred in 15% of participants in the active group of a phase II trial (Hajós et al., 2024) and was linked to worsened pre-existing tinnitus in one feasibility study participant after a single session (He et al., 2021). However, in our study, no tinnitus was observed, even in a participant with a history of the condition. Lastly, the safety results from this study suggest that strategies like implementing a gradual off-boarding process to help participants readjust to their surroundings and using VR-specific lenses to reduce visual strain could effectively mitigate transient side effects such as disorientation and blurred vision in future applications.

Efficacy-wise, this study demonstrated that VR-based GSS can drive gamma activity, with effects observed across different sensory modalities, stimulus complexities, and tasks with varying cognitive demands. Both visual and auditory stimulation independently induced significant gamma oscillatory activity, as evidenced by significant fold changes in power from baseline within corresponding sensory cortical regions (Figure 1). Importantly, the study also highlighted the design flexibility of VR-based GSS since comparing simple stimuli, such as black-and-white flickers and 40Hz clicks, with complex stimuli, including bright videos and amplitude-modulated pure tones, revealed no significant differences in gamma power increase (Figure 1B). This suggests that engaging, contextually-rich content can be paired with gamma stimulation without significantly diminishing its efficacy, offering exciting opportunities for personalized, user-friendly interventions that promote adherence and therapeutic engagement.

When combining auditory and visual 40Hz stimuli, we evaluated gamma activity modulation under both controlled and naturalistic conditions, employing passive as well as cognitively engaging GSS protocols. Controlled scenarios employed repetitive multimedia content presented with and without 40Hz modulation, while naturalistic conditions involved varied 40Hz modulated and unmodulated audiovisual pairs in an associative memory task. Across both paradigms, our VR-based GSS approach reliably modulated gamma neural activity, evidenced by significant increases in both power and temporal alignment during the active condition compared to the sham (Figures 2-4). While our protocol was not powered to systematically compare the effects of audiovisual material flickerd at 40Hz with varying cognitive loads on the topography of gamma modulation, it highlights an important avenue for future research.

Behavioral effects of non-invasive neuromodulation interventions are generally expected to emerge from prolonged exposure rather than a single session (Chan et al., 2022; C. Liu et al., 2022; Nissim et al., 2023). Nonetheless, given the critical role of gamma synchrony in binding information across brain regions, and its association with the precise temporal coordination of neuronal assemblies involved in encoding (Nyhus & Curran, 2010), we explored whether modulating the volume and brightness of video-sound pairs at 40Hz could facilitate memory formation. Despite finding no significant differences in recall accuracy between modulated and unmodulated trials in our associative memory task, further analyses are warranted. Specifically, and consistent with previous research (Wang et al., 2018, 2023), assessing the strength of evoked gamma synchrony between auditory and visual cortices and its relationship to memory performance will be crucial for testing the hypothesis that stronger gamma synchrony induced by GSS enhances the integration of visual and auditory streams, ultimately facilitating memory formation.

Overall, our feasibility findings validate VR technology as a flexible and versatile neuromodulation tool capable of delivering auditory and visual sensory stimulation independently or in combination, embedded in diverse multimedia content and during passive or cognitively active states. The importance of such flexibility lies in the ability to strategically embed stimuli in contextually relevant content, which previous studies suggest can shape the propagation and magnitude of neural entrainment to sensory stimulation. For instance, in a study by Khachatryan et al., 2022, EEG recordings from 15 healthy young participants during 40Hz visual stimulation showed that the topography of entrained electrodes during 40Hz visual stimulation varied significantly depending on the cognitive task paired with the stimulation—mental counting predominantly activated parietocentral regions, while visual attention tasks engaged centro-frontal areas (Khachatryan et al., 2022). Similarly, sensory stimulation delivered at 5Hz to six drug-resistant epileptic patients entrained deep brain regions, such as the hippocampus and the amygdala, only when the emotional valence of the flickered stimuli was non-neutral, and not when it was neutral (Hoyer et al., 2025). Crucially, the same study found that stimuli valence influenced not only the topography of gamma modulation and but also behavioral outcomes. Specifically, in a complementary experiment involving 27 healthy participants, the study reported that negative valence stimulation, but not stimulation with a neutral valence, selectively enhanced recognition memory for target items encountered during the stimulation period compared to distractor items (Hoyer et al., 2025). These findings highlight the potential of incorporating contextually rich content into sensory stimulation to target specific brain networks linked to particular symptoms of AD. By leveraging these insights, VR-based GSS stands as a promising platform for advancing precision medicine in AD care, offering the potential to deliver the neuroprotective and synaptic plasticity-enhancing benefits of GSS to specific brain regions, effectively addressing distinct, patient-specific cognitive and functional deficits, and optimizing therapeutic outcomes.

Whether VR-based GSS would be well-accepted for daily at-home use by an AD population remains to be tested in future longitudinal studies; however, results from this pilot study are encouraging. While older adults are often thought to be less receptive to advanced technologies like VR, our findings challenge this notion, with ∼94% of participants describing the VR experience as non-overwhelming. Additionally, despite the repetitive nature of the three experimental tasks which averaged to a total approximate duration of 1h30 and which did not represent a fully optimized therapeutic protocol, more than half of the participants (∼63%) rated the experience as enjoyable. These results align with studies showing that VR-based interventions can enhance cognitive engagement and emotional well-being in older populations, even among individuals with subjective or objective cognitive impairments (Arlati et al., 2021; Bauer & Andringa, 2020). Furthermore and in support of the VR task-based GSS approach proposed above, immersive VR environments have also been shown to offer meaningful cognitive training and rehabilitation experiences, with users reporting positive emotional responses and minimal adverse effects (Wais et al., 2021; Ziegler et al., 2022).

Lastly, the complete absence of motion sickness symptoms in our cohort is of particular importance, given its prevalence in VR applications (Chattha et al., 2020). This favorable outcome likely stems from deliberate design and hardware choices, including presenting 40Hz-modulated content in 2D to create a stable, theater-like experience, and utilizing low-persistence organic light-emitting diode (OLED) screens that minimize motion blur and enhance visual clarity.

### Limitations

While this study provides a critical proof of concept for VR-based gamma sensory stimulation (GSS), a few limitations should be acknowledged. As gamma sensory stimulation gains recognition for its therapeutic potential, there is increasing interest in translating this method into accessible, user-friendly solutions. However, rigorous testing of the technology used is essential to ensure therapeutic safety and efficacy. VR headsets, originally designed for interactive gaming and entertainment, are not inherently optimized for precision neural stimulation. While our experimental set up did not allow for simultaneous acquisition of stimuli and brain data, we systematically evaluated the performance of off-the-shelf VR devices using Arduino-based sound and light sensors before and after each session. These tests demonstrated that the VR-based GSS protocol used in this study could reliably deliver 40Hz stimulation as evidenced by the average PSD peak of 39.95 ± 0.67 Hz and 40.43 ± 0.52 Hz for visual and auditory stimuli, respectively. However, phase synchrony analysis comparing a simulated 40Hz sinusoidal waveform to visual stimulation trials recorded with a photodiode indicated relatively low stability in the visual stimulation (PLV = 0.65 ± 0.16). While this did not hinder our stimulation approach from driving gamma neural activity in response to visual 40Hz stimulation (Figure 1 A and B), these findings highlight opportunities to further refine VR technology for neuromodulation applications.

In addition, following the positive results of this pilot study, the safety, efficacy, and tolerability of VR-based GSS must be investigated longitudinally in a double-blind and sham controlled study and in a diverse cohort of people with AD.

## Conclusion

This single-day study demonstrates that VR technology can effectively deliver gamma sensory stimulation (GSS). Using 40Hz-modulated audiovisual content, both passive and cognitively engaging conditions successfully and safely modulated gamma-frequency neural activity. The flexibility of this approach supports the development of engaging and potentially more effective GSS protocols, paving the way for scalable and personalized therapies to address the unmet needs of Alzheimer’s disease and other neurodegenerative disorders.

## Authors contribution

Conceptualization: C.R., T.Z., and A.C.; Methodology: C.R., H.A., A.C., and S.H.; Software: G.H. and C.R.; Formal Analysis: H.A. and C.R.; Investigation: G.H., S.N., and C.R.; Resources: R.C.; Data Curation: G.H. and H.A.; Writing – Original Draft: C.R. and H.A.; Writing – Review and Editing: H.A., G.H., S.N., R.C., T.Z., A.C., and S.H.; Supervision: C.R.

## Data Availability

All data produced in the present study are available upon reasonable request to the authors.

## Acknowledgements

We extend our sincere gratitude to Sue Fairley and all the residents at The Sequoias Portola Valley who participated in the study for their time and support. We also thank Maxime Ronceray, Khadijeh Sadatnejad, and Timothy Nolan for their contributions to the development of this protocol and their technical support. This work was supported by funding from Région Île-de-France, France 2030, and Bpifrance.

## References

1. Arlati, S., Di Santo, S. G., Franchini, F., Mondellini, M., Filiputti, B., Luchi, M., Ratto, F., Ferrigno, G., Sacco, M., & Greci, L. (2021). Acceptance and Usability of Immersive Virtual Reality in Older Adults with Objective and Subjective Cognitive Decline. Journal of Alzheimer’s Disease: JAD, 80(3), 1025–1038. 10.3233/JAD-201431

2. Arnal, L. H., Kleinschmidt, A., Spinelli, L., Giraud, A.-L., & Mégevand, P. (2019). The rough sound of salience enhances aversion through neural synchronisation. Nature Communications, 10(1), 3671. 10.1038/s41467-019-11626-7

3. Bartoli, E., Bosking, W., Chen, Y., Li, Y., Sheth, S. A., Beauchamp, M. S., Yoshor, D., & Foster, B. L. (2019). Functionally Distinct Gamma Range Activity Revealed by Stimulus Tuning in Human Visual Cortex. Current Biology : CB, 29(20), 3345–3358.e7. 10.1016/j.cub.2019.08.004

4. Bauer, A. C. M., & Andringa, G. (2020). The Potential of Immersive Virtual Reality for Cognitive Training in Elderly. Gerontology, 66(6), 614–623. 10.1159/000509830

5. Benussi, A., Cantoni, V., Grassi, M., Brechet, L., Michel, C. M., Datta, A., Thomas, C., Gazzina, S., Cotelli, M. S., Bianchi, M., Premi, E., Gadola, Y., Cotelli, M., Pengo, M., Perrone, F., Scolaro, M., Archetti, S., Solje, E., Padovani, A., … Borroni, B. (2022). Increasing Brain Gamma Activity Improves Episodic Memory and Restores Cholinergic Dysfunction in Alzheimer’s Disease. Annals of Neurology, 92(2), 322–334. 10.1002/ana.26411

6. Blanpain, L. T., Cole, E. R., Chen, E., Park, J. K., Walelign, M. Y., Gross, R. E., Cabaniss, B. T., Willie, J. T., & Singer, A. C. (2024). Multisensory flicker modulates widespread brain networks and reduces interictal epileptiform discharges. Nature Communications, 15(1), 3156. 10.1038/s41467-024-47263-y

7. Chan, D., Suk, H.-J., Jackson, B. L., Milman, N. P., Stark, D., Klerman, E. B., Kitchener, E., Fernandez Avalos, V. S., de Weck, G., Banerjee, A., Beach, S. D., Blanchard, J., Stearns, C., Boes, A. D., Uitermarkt, B., Gander, P., Howard, M., Sternberg, E. J., Nieto-Castanon, A., … Tsai, L.-H. (2022). Gamma frequency sensory stimulation in mild probable Alzheimer’s dementia patients: Results of feasibility and pilot studies. PloS One, 17(12), e0278412. 10.1371/journal.pone.0278412

8. Chattha, U. A., Janjua, U. I., Anwar, F., Madni, T. M., Cheema, M. F., & Janjua, S. I. (2020). Motion Sickness in Virtual Reality: An Empirical Evaluation. IEEE Access, 8, 130486– 130499. IEEE Access. 10.1109/ACCESS.2020.3007076

9. Cimenser, A., Hempel, E., Travers, T., Strozewski, N., Martin, K., Malchano, Z., & Hajós, M. (2021). Sensory-Evoked 40-Hz Gamma Oscillation Improves Sleep and Daily Living Activities in Alzheimer’s Disease Patients. Frontiers in Systems Neuroscience, 15, 746859. 10.3389/fnsys.2021.746859

10. Clouter, A., Shapiro, K. L., & Hanslmayr, S. (2017). Theta Phase Synchronization Is the Glue that Binds Human Associative Memory. Current Biology, 27(20), 3143–3148.e6. 10.1016/j.cub.2017.09.001

11. Crews, L., & Masliah, E. (2010). Molecular mechanisms of neurodegeneration in Alzheimer’s disease. Human Molecular Genetics, 19(R1), R12–R20. 10.1093/hmg/ddq160

12. Da, X., Hempel, E., Ou, Y., Rowe, O. E., Malchano, Z., Hajós, M., Kern, R., Megerian, J. T., & Cimenser, A. (2024). Noninvasive Gamma Sensory Stimulation May Reduce White Matter and Myelin Loss in Alzheimer’s Disease. Journal of Alzheimer’s Disease: JAD, 97(1), 359–372. 10.3233/JAD-230506

13. Duecker, K., Gutteling, T. P., Herrmann, C. S., & Jensen, O. (2021). No Evidence for Entrainment: Endogenous Gamma Oscillations and Rhythmic Flicker Responses Coexist in Visual Cortex. The Journal of Neuroscience, 41(31), 6684–6698. 10.1523/JNEUROSCI.3134-20.2021

14. Fries, P. (2015). Rhythms For Cognition: Communication Through Coherence. Neuron, 88(1), 220–235. 10.1016/j.neuron.2015.09.034

15. Gramfort, A., Luessi, M., Larson, E., Engemann, D. A., Strohmeier, D., Brodbeck, C., Goj, R., Jas, M., Brooks, T., Parkkonen, L., & Hämäläinen, M. (2013). MEG and EEG data analysis with MNE-Python. Frontiers in Neuroscience, 7. 10.3389/fnins.2013.00267

16. Hajós, M., Boasso, A., Hempel, E., Shpokayte, M., Konisky, A., Kwan, K., Hendrix, S., Megerian, J. T., & Malchano, Z. (2024). Safety, tolerability, and efficacy estimate of evoked gamma oscillation in mild to moderate Alzheimer’s disease. Frontiers in Neurology, 15. 10.3389/fneur.2024.1343588

17. Hanslmayr, S., Axmacher, N., & Inman, C. S. (2019). Modulating Human Memory via Entrainment of Brain Oscillations. Trends in Neurosciences, 42(7), 485–499. 10.1016/j.tins.2019.04.004

18. Headley, D. B., & Weinberger, N. M. (2011). Gamma-Band Activation Predicts Both Associative Memory and Cortical Plasticity. Journal of Neuroscience, 31(36), 12748–12758. 10.1523/JNEUROSCI.2528-11.2011

19. Hoyer, R. S., Bernier, M.-F., Martineau, L., Bonaventure, P. L., Labelle, C., Borderie, A., Blanchette, I., Blumenthal, A., & Albouy, P. (2025). Valence-dependent sensory-rhythmic neural entrainment modulates cortico-subcortical dynamics, attention, and memory (p. 2025.01.03.631218). bioRxiv. 10.1101/2025.01.03.631218

20. Iaccarino, H. F., Singer, A. C., Martorell, A. J., Rudenko, A., Gao, F., Gillingham, T. Z., Mathys, H., Seo, J., Kritskiy, O., Abdurrob, F., Adaikkan, C., Canter, R. G., Rueda, R., Brown, E. N., Boyden, E. S., & Tsai, L.-H. (2016). Gamma frequency entrainment attenuates amyloid load and modifies microglia. Nature, 540(7632), 230–235. 10.1038/nature20587

21. Jensen, O., Kaiser, J., & Lachaux, J.-P. (2007). Human gamma-frequency oscillations associated with attention and memory. Trends in Neurosciences, 30(7), 317–324. 10.1016/j.tins.2007.05.001

22. Jones, K. T., Ostrand, A. E., Gazzaley, A., & Zanto, T. P. (2023). Enhancing cognitive control in amnestic mild cognitive impairment via at-home non-invasive neuromodulation in a randomized trial. Scientific Reports, 13(1), 7435. 10.1038/s41598-023-34582-1

23. Jones, M., McDermott, B., Oliveira, B. L., O’Brien, A., Coogan, D., Lang, M., Moriarty, N., Dowd, E., Quinlan, L., Mc Ginley, B., Dunne, E., Newell, D., Porter, E., Elahi, M. A., O’ Halloran, M., & Shahzad, A. (2019). Gamma Band Light Stimulation in Human Case Studies: Groundwork for Potential Alzheimer’s Disease Treatment. Journal of Alzheimer’s Disease: JAD, 70(1), 171–185. 10.3233/JAD-190299

24. Jung, Y. H., Jang, H., Park, S., Kim, H. J., Seo, S. W., Kim, G. B., Shon, Y.-M., Kim, S., & Na, D. L. (2024). Effectiveness of Personalized Hippocampal Network–Targeted Stimulation in Alzheimer Disease: A Randomized Clinical Trial. JAMA Network Open, 7(5), e249220. 10.1001/jamanetworkopen.2024.9220

25. Khachatryan, E., Wittevrongel, B., Reinartz, M., Dauwe, I., Carrette, E., Meurs, A., Van Roost, D., Boon, P., & Van Hulle, M. M. (2022). Cognitive tasks propagate the neural entrainment in response to a visual 40 Hz stimulation in humans. Frontiers in Aging Neuroscience, 14, 1010765. 10.3389/fnagi.2022.1010765

26. Koch, G., Casula, E. P., Bonnì, S., Borghi, I., Assogna, M., Minei, M., Pellicciari, M. C., Motta, C., D’Acunto, A., Porrazzini, F., Maiella, M., Ferrari, C., Caltagirone, C., Santarnecchi, E., Bozzali, M., & Martorana, A. (2022). Precuneus magnetic stimulation for Alzheimer’s disease: A randomized, sham-controlled trial. Brain, 145(11), 3776–3786. 10.1093/brain/awac285

27. Kurimoto, R., Ishii, R., Canuet, L., Ikezawa, K., Iwase, M., Azechi, M., Aoki, Y., Ikeda, S., Yoshida, T., Takahashi, H., Nakahachi, T., Kazui, H., & Takeda, M. (2012). Induced oscillatory responses during the Sternberg’s visual memory task in patients with Alzheimer’s disease and mild cognitive impairment. NeuroImage, 59(4), 4132–4140. 10.1016/j.neuroimage.2011.10.061

28. Lachaux, J.-P., George, N., Tallon-Baudry, C., Martinerie, J., Hugueville, L., Minotti, L., Kahane, P., & Renault, B. (2005). The many faces of the gamma band response to complex visual stimuli. NeuroImage, 25(2), 491–501. 10.1016/j.neuroimage.2004.11.052

29. Lahijanian, M., Aghajan, H., & Vahabi, Z. (2024). Auditory gamma-band entrainment enhances default mode network connectivity in dementia patients. Scientific Reports, 14(1), 13153. 10.1038/s41598-024-63727-z

30. Liu, C., Han, T., Xu, Z., Liu, J., Zhang, M., Du, J., Zhou, Q., Duan, Y., Li, Y., Wang, J., Cui, D., & Wang, Y. (2022). Modulating Gamma Oscillations Promotes Brain Connectivity to Improve Cognitive Impairment. Cerebral Cortex (New York, N.Y.: 1991), 32(12), 2644–2656. 10.1093/cercor/bhab371

31. Liu, Q., Contreras, A., Afaq, M. S., Yang, W., Hsu, D. K., Russell, M., Lyeth, B., Zanto, T. P., & Zhao, M. (2023). Intensity-dependent gamma electrical stimulation regulates microglial activation, reduces beta-amyloid load, and facilitates memory in a mouse model of Alzheimer’s disease. Cell & Bioscience, 13(1), 138. 10.1186/s13578-023-01085-5

32. Mably, A. J., & Colgin, L. L. (2018). Gamma oscillations in cognitive disorders. Current Opinion in Neurobiology, 52, 182–187. 10.1016/j.conb.2018.07.009

33. Martorell, A. J., Paulson, A. L., Suk, H.-J., Abdurrob, F., Drummond, G. T., Guan, W., Young, J. Z., Kim, D. N.-W., Kritskiy, O., Barker, S. J., Mangena, V., Prince, S. M., Brown, E. N., Chung, K., Boyden, E. S., Singer, A. C., & Tsai, L.-H. (2019). Multi-sensory Gamma Stimulation Ameliorates Alzheimer’s-Associated Pathology and Improves Cognition. Cell, 177(2), 256–271.e22. 10.1016/j.cell.2019.02.014

34. Murdock, M. H., Yang, C.-Y., Sun, N., Pao, P.-C., Blanco-Duque, C., Kahn, M. C., Kim, T., Lavoie, N. S., Victor, M. B., Islam, M. R., Galiana, F., Leary, N., Wang, S., Bubnys, A., Ma, E., Akay, L. A., Sneve, M., Qian, Y., Lai, C., … Tsai, L.-H. (2024). Multisensory gamma stimulation promotes glymphatic clearance of amyloid. Nature, 627(8002), 149–156. 10.1038/s41586-024-07132-6

35. Nissim, N. R., Pham, D. V. H., Poddar, T., Blutt, E., & Hamilton, R. H. (2023). The impact of gamma transcranial alternating current stimulation (tACS) on cognitive and memory processes in patients with mild cognitive impairment or Alzheimer’s disease: A literature review. Brain Stimulation, 16(3), 748–755. 10.1016/j.brs.2023.04.001

36. Nyhus, E., & Curran, T. (2010). Functional role of gamma and theta oscillations in episodic memory. Neuroscience & Biobehavioral Reviews, 34(7), 1023–1035. 10.1016/j.neubiorev.2009.12.014

37. Pileckyte, I., & Soto-Faraco, S. (2024). Sensory stimulation enhances visual working memory capacity. Communications Psychology, 2(1), 1–18. 10.1038/s44271-024-00158-6

38. Sims, J. R., Zimmer, J. A., Evans, C. D., Lu, M., Ardayfio, P., Sparks, J., Wessels, A. M., Shcherbinin, S., Wang, H., Monkul Nery, E. S., Collins, E. C., Solomon, P., Salloway, S., Apostolova, L. G., Hansson, O., Ritchie, C., Brooks, D. A., Mintun, M., Skovronsky, D. M., & TRAILBLAZER-ALZ 2 Investigators. (2023). Donanemab in Early Symptomatic Alzheimer Disease: The TRAILBLAZER-ALZ 2 Randomized Clinical Trial. JAMA, 330(6), 512–527. 10.1001/jama.2023.13239

39. Singh, D. (2022). Astrocytic and microglial cells as the modulators of neuroinflammation in Alzheimer’s disease. Journal of Neuroinflammation, 19(1), Article 1. 10.1186/s12974-022-02565-0

40. Tahami Monfared, A. A., Byrnes, M. J., White, L. A., & Zhang, Q. (2022). The Humanistic and Economic Burden of Alzheimer’s Disease. Neurology and Therapy, 11(2), 525–551. 10.1007/s40120-022-00335-x

41. Tallon-Baudry, C., Bertrand, O., Peronnet, F., & Pernier, J. (1998). Induced γ-Band Activity during the Delay of a Visual Short-Term Memory Task in Humans. Journal of Neuroscience, 18(11), 4244–4254. 10.1523/JNEUROSCI.18-11-04244.1998

42. Tarasoff-Conway, J. M., Carare, R. O., Osorio, R. S., Glodzik, L., Butler, T., Fieremans, E., Axel, L., Rusinek, H., Nicholson, C., Zlokovic, B. V., Frangione, B., Blennow, K., Ménard, J., Zetterberg, H., Wisniewski, T., & de Leon, M. J. (2015). Clearance systems in the brain-implications for Alzheimer disease. Nature Reviews. Neurology, 11(8), 457–470. 10.1038/nrneurol.2015.119

43. van Dyck, C. H., Swanson, C. J., Aisen, P., Bateman, R. J., Chen, C., Gee, M., Kanekiyo, M., Li, D., Reyderman, L., Cohen, S., Froelich, L., Katayama, S., Sabbagh, M., Vellas, B., Watson, D., Dhadda, S., Irizarry, M., Kramer, L. D., & Iwatsubo, T. (2023). Lecanemab in Early Alzheimer’s Disease. New England Journal of Medicine, 388(1), 9–21. 10.1056/NEJMoa2212948

44. Wais, P. E., Arioli, M., Anguera-Singla, R., & Gazzaley, A. (2021). Virtual reality video game improves high-fidelity memory in older adults. Scientific Reports, 11(1), Article 1. 10.1038/s41598-021-82109-3

45. Wang, D., Clouter, A., Chen, Q., Shapiro, K. L., & Hanslmayr, S. (2018). Single-Trial Phase Entrainment of Theta Oscillations in Sensory Regions Predicts Human Associative Memory Performance. Journal of Neuroscience, 38(28), 6299–6309. 10.1523/JNEUROSCI.0349-18.2018

46. Wang, D., Shapiro, K. L., & Hanslmayr, S. (2023). Altering stimulus timing via fast rhythmic sensory stimulation induces STDP-like recall performance in human episodic memory. Current Biology, 33(15), 3279–3288.e7. 10.1016/j.cub.2023.06.062

47. Williams, S. D., Setzer, B., Fultz, N. E., Valdiviezo, Z., Tacugue, N., Diamandis, Z., & Lewis, L. D. (2023). Neural activity induced by sensory stimulation can drive large-scale cerebrospinal fluid flow during wakefulness in humans. PLOS Biology, 21(3). 10.1371/journal.pbio.3002035

48. Ziegler, D. A., Anguera, J. A., Gallen, C. L., Hsu, W.-Y., Wais, P. E., & Gazzaley, A. (2022). Leveraging technology to personalize cognitive enhancement methods in aging. Nature Aging, 2(6), Article 6. 10.1038/s43587-022-00237-5

